# Optimal environmental testing frequency for outbreak surveillance

**DOI:** 10.1101/2023.09.14.23295550

**Authors:** Jason W. Olejarz, Kirstin I. Oliveira Roster, Stephen M. Kissler, Marc Lipsitch, Yonatan H. Grad

**Affiliations:** Department of Immunology and Infectious Diseases, Harvard T. H. Chan School of Public Health, Boston, MA 02115, USA; Center for Communicable Disease Dynamics, Harvard T. H. Chan School of Public Health, Boston, MA 02115, USA; Department of Computer Science, University of Colorado Boulder, Boulder, CO 80309, USA; Department of Epidemiology, Harvard T. H. Chan School of Public Health, Boston, MA 02115, USA

**Keywords:** Environmental surveillance, Early pathogen detection, Wastewater sampling, Vector trapping, Mathematical modeling

## Abstract

Public health surveillance for pathogens presents an optimization problem: we require enough sampling to identify intervention-triggering shifts in pathogen epidemiology, such as new introductions or sudden increases in prevalence, but not so much that costs due to surveillance itself outweigh those from pathogen-associated illness. To determine this optimal sampling frequency, we developed a general mathematical model for the introduction of a new pathogen that, once introduced, increases in prevalence exponentially. Given the relative cost of infection *vs*. sampling, we derived equations for the expected combined cost of disease burden and surveillance given a sampling frequency and thus the sampling frequency for which the expected total cost is lowest.

## Introduction

A key goal of public health infectious disease surveillance systems is to detect a pathogen at an early stage of its entry into the population, enabling interventions to limit its spread and the harm it could inflict [1, 2, 3]. Such efforts are increasingly important given the many ways in which communities are connected, with growing populations, global travel, and urbanization, and given ecological shifts associated with climate change and other factors leading to emergence and re-emergence of vector-borne diseases, with cases of locally acquired dengue and malaria where they had been absent for many decades [4, 5, 6].

One important strategy for achieving early pathogen detection is monitoring for infected individuals through robust clinical surveillance. However, clinical surveillance is inherently limited in important ways. Infections may be mildly symptomatic, asymptomatic, or have a long pre-symptomatic infectious phase, such that the pathogen population has spread extensively before the first clinical cases are diagnosed and the pathogen identified [7]. In contexts where access to care or resources are limited, missed cases and reporting delays can make it difficult to rapidly detect and correctly diagnose new infections [8].

For pathogens that can be detected in environmental samples and that spread by vectors, a complementary and critical strategy for early pathogen detection is monitoring through periodic sampling of the environment. Pathogen detection in wastewater has been important for the surveillance and control of poliovirus [9, 10] and has been used more recently for tracking the local epidemic dynamics and evolution of SARS-CoV-2 [11, 12], norovirus [13], influenza [14], mpox [15, 16], and other pathogens [17]. Efforts are underway to extend these techniques for tracking antibiotic resistance genes in wastewater [18, 19]. For vector-borne pathogens, including West Nile virus [20], *Borrelia* species [21], and Powassan virus [22], surveillance includes pathogen detection in vectors collected via traps, with sampling also taking place at a given frequency.

Monitoring for infectious diseases requires substantial time, money, and infrastructure for detection, interpretation, and response [23, 24, 25, 26, 27, 28]. Although the potential for environmental and vector-based surveillance systems have been recognized and widely discussed, and despite the massive push to fund and develop these programs, particularly wastewater efforts [29, 30], there remains a critical gap in our understanding: How should surveillance be designed to achieve maximal effectiveness [31, 32, 33, 34, 35, 36]? A central consideration is how often testing should be performed (Figure 1). Here, we addressed this question by formulating a simple, stochastic model for pathogen introduction, growth, and detection in the presence of periodic sampling and testing. We identified the key parameters of this process, and we derived a simple equation for the expected total cost (i.e., the sum of all costs related to surveillance and to effects from the disease, when considered as an average over many realizations of the stochastic dynamics). The expected total cost is a function of the parameters of the model, and given values for these parameters, we can minimize the expected total cost.

**Figure 1:**
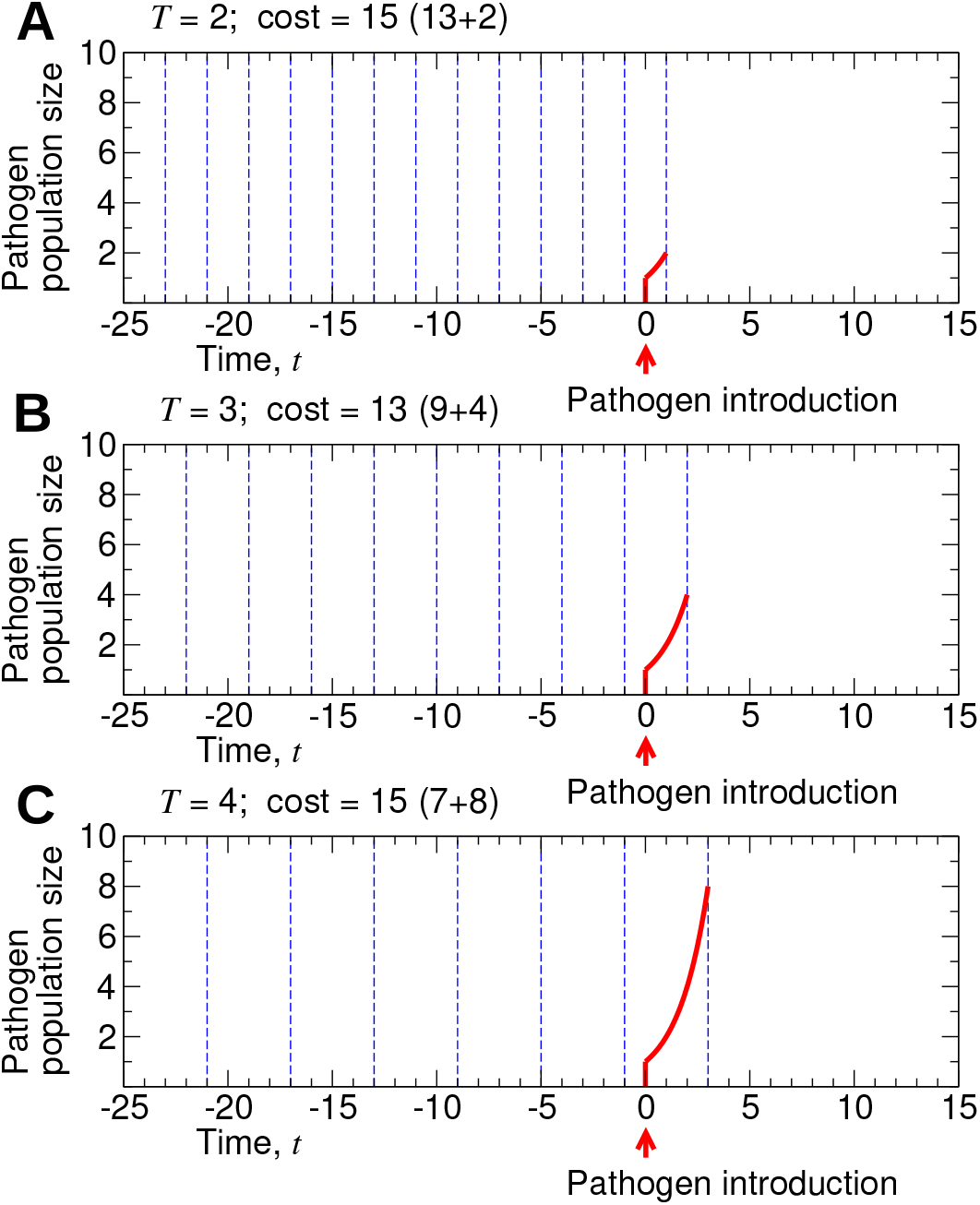
Optimization of surveillance. A simple scenario illustrates how surveillance can be optimized. In the plot, the dotted blue lines represent sampling events, and the solid red curves represent the abundance of a pathogen. Here, we assume that a pathogen first emerges at time *t* = 0 with the pathogen population growing exponentially, doubling at each subsequent time step. Sampling of the environment occurs at times −25 + *nT* —where *T* is the sampling period and *n* ≥ 1 is an integer—until the pathogen is first detected. We plot the outcomes if the sampling period had been (A) *T* = 2, (B) *T* = 3, or (C) *T* = 4. If the cost associated with one sampling event is equal to the cost associated with one instance of the pathogen, and if costs accumulate linearly, then *T* = 3 would have resulted in the lowest total cost.

Our goal was to minimize the expected total surveillance and disease cost for the detection of the first appearance of a pathogen. Accordingly, we employed a simple model of surveillance to detect the entry of a pathogen into a population, assuming that, once the pathogen is present, its prevalence increases exponentially. Sampling begins at time *t* = 0 and continues regularly at time *t*_*n*_ = *nT*, where *n* ≥ 1 and *T* is the sampling period. Each sampling event incurs a cost *c*_1_, and since there are 1*/T* sampling events per unit time, the sampling cost per unit time is given by *c*_1_*/T* . Copies of the pathogen are introduced after time *t* = 0 according to a Poisson process, such that the waiting times between initiation events are exponentially distributed with rate *λ*. Once a new lineage is introduced, it also reproduces (transmits) according to a Poisson process, such that the expected prevalence of the pathogen grows exponentially with rate *r*. If a sampling event detects a copy of a pathogen that belongs to a particular lineage, then that lineage is “detected.” Let *p* be the probability that a sampling event detects a copy of a pathogen, and since each detection occurs independently, the probability that a sampling event detects a lineage of size *n* is given by 1 − (1 − *p*)^*n*^. Once a lineage is detected, we assume that intervention is immediate and is successful at suppressing further spread of that lineage. If a lineage has *N* copies of the pathogen when it is detected, then the disease cost due to that lineage is given by *c*_2_*N* . Letting ⟨*N* ⟩ denote the expected size of a lineage when it is detected, the expected disease cost due to a lineage is given by *c*_2_⟨*N* ⟩. Since new lineages appear at rate *λ*, the expected disease cost per unit time is given by *λc*_2_⟨*N* ⟩. The expected total cost per unit time is then *c*_1_*/T* + *λc*_2_⟨*N* ⟩. The model is illustrated in Figure 2.

**Figure 2:**
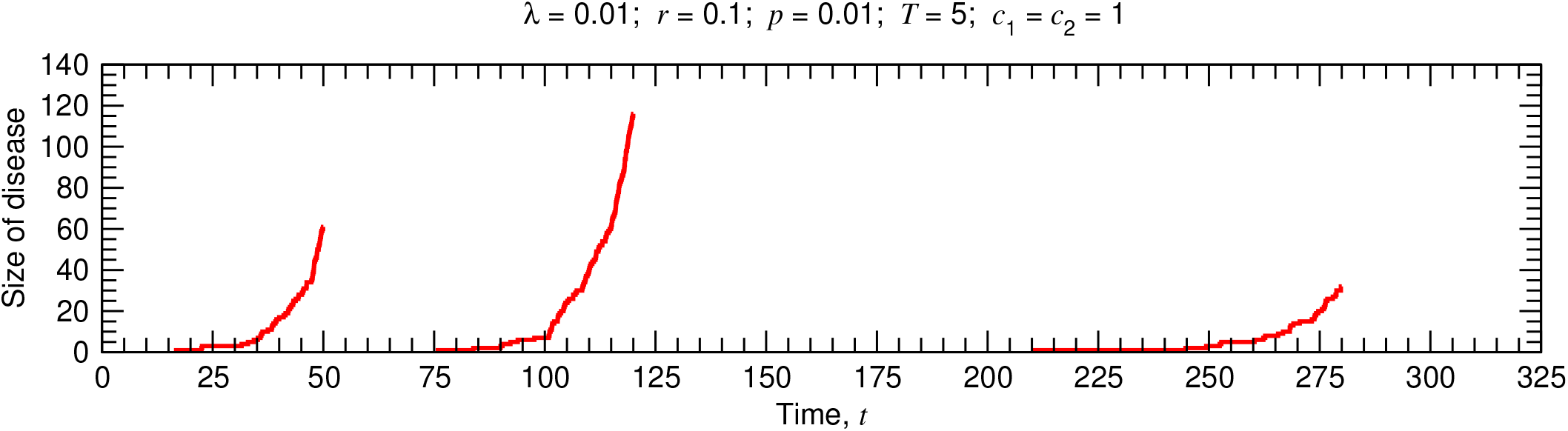
Stochastic surveillance model. A single realization of the stochastic surveillance and disease dynamics is shown. The environment is tested at times 5*n*, where *n* is an integer and 0 ≤ *n* ≤ 65. There are thus 66 testing events, so the cumulative surveillance cost is 66. The first lineage begins at time *t* ≈ 16.7 and is detected at time *t* = 50, when its size is 61. The second lineage begins at time *t* ≈ 75.8 and is detected at time *t* = 120, when its size is 116. The third lineage begins at time *t* ≈ 210.6 and is detected at time *t* = 280, when its size is 32. The cumulative disease cost is thus 61 + 116 + 32 = 209. The total surveillance and disease cost is 66 + 209 = 275, and the total cost per unit time is 275*/*325 = 11*/*13.

## Results

We derived an accurate approximation for the expected total cost of testing and disease burden per unit time, ⟨*C*⟩:

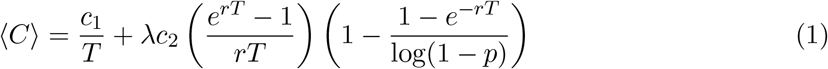

Details on the derivation of Equation (1) are provided in the Supplementary Information. By comparing Equation (1) with *c*_1_*/T* + *λc*_2_⟨N ⟩, notice that the product of the two factors in large parentheses is equal to the expected size of an outbreak when it is detected, ⟨N ⟩. It is helpful to understand the behavior of ⟨N ⟩ as a function of *p, r*, and *T* . In the limit of a perfectly sensitive detector (i.e., *p* → 1), we have − log(1 − *p*) → ∞, and the second factor in big parentheses just approaches 1. So as *p* → 1, we have ⟨N ⟩ ≈ (*e*^*rT*^ − 1)*/*(*rT* ), which is the expected size of the outbreak when the first test happens, given an introduction. If the detector has poor sensitivity (i.e., *p* ≪ 1), then − log(1 − *p*) ≈ *p*, and the second factor in big parentheses is approximately equal to (1 − *e*^−*rT*^ )*/p*. So for small values of *p*, we have ⟨N ⟩ ≈ (*e*^*rT*^ − 1)(1 − *e*^−*rT*^ )*/*(*rTp*). For large values of the pathogen growth rate, *r*, we have ⟨N ⟩ ≈ [*e*^*rT*^ */*(*rT* )][1 − 1*/* log(1 − *p*)], and for small values of *r*, we have ⟨N ⟩ → 1—i.e., the pathogen is detected before it has a chance to produce new infections. Since ⟨N ⟩ is a function of the product *rT* (not of *r* and *T* individually), its behavior as a function of the testing period, *T*, is the same: For large values of *T*, we have ⟨N ⟩ ≈ [*e*^*rT*^ */*(*rT* )][1 −1*/* log(1 −*p*)], and for small values of *T*, we have ⟨N ⟩ → 1. Notice that ⟨N ⟩ is a decreasing function of *p*, an increasing function of *r*, and an increasing function of *T* —i.e., a less-sensitive detector, a more rapidly growing pathogen, or a larger testing interval all result in a larger expected size of the outbreak when it is detected.

The expected infection cost per unit time, *λc*_2_⟨N ⟩, is therefore also an increasing function of the testing period, *T*, and this quantity becomes arbitrarily large as *T* → ∞. The surveillance cost per unit time, *c*_1_*/T*, however, is a decreasing function of *T*, and this quantity becomes arbitrarily large as *T* → 0. These behaviors are evident in Figure 3, where we plotted (*C*) as a function of the sampling frequency, *f* = 1*/T*, for different values of the five model parameters. It is instructive to consider the effects of very large or very small values of *f* on (*C*). As we increase *f*, we can detect the disease more rapidly, thereby diminishing disease-related costs. But returns are diminishing: We can—at best—hope to discover the disease as a single unit, representing the pathogen introduction, before it has begun to spread and proliferate, while increasing *f* further can add arbitrarily large surveillance costs. At the other extreme, setting *f* too small allows the disease to proliferate before any intervention is applied. Therefore, as shown in each of the curves in Figure 3, (*C*) attains a minimum for a particular value of *f* .

**Figure 3:**
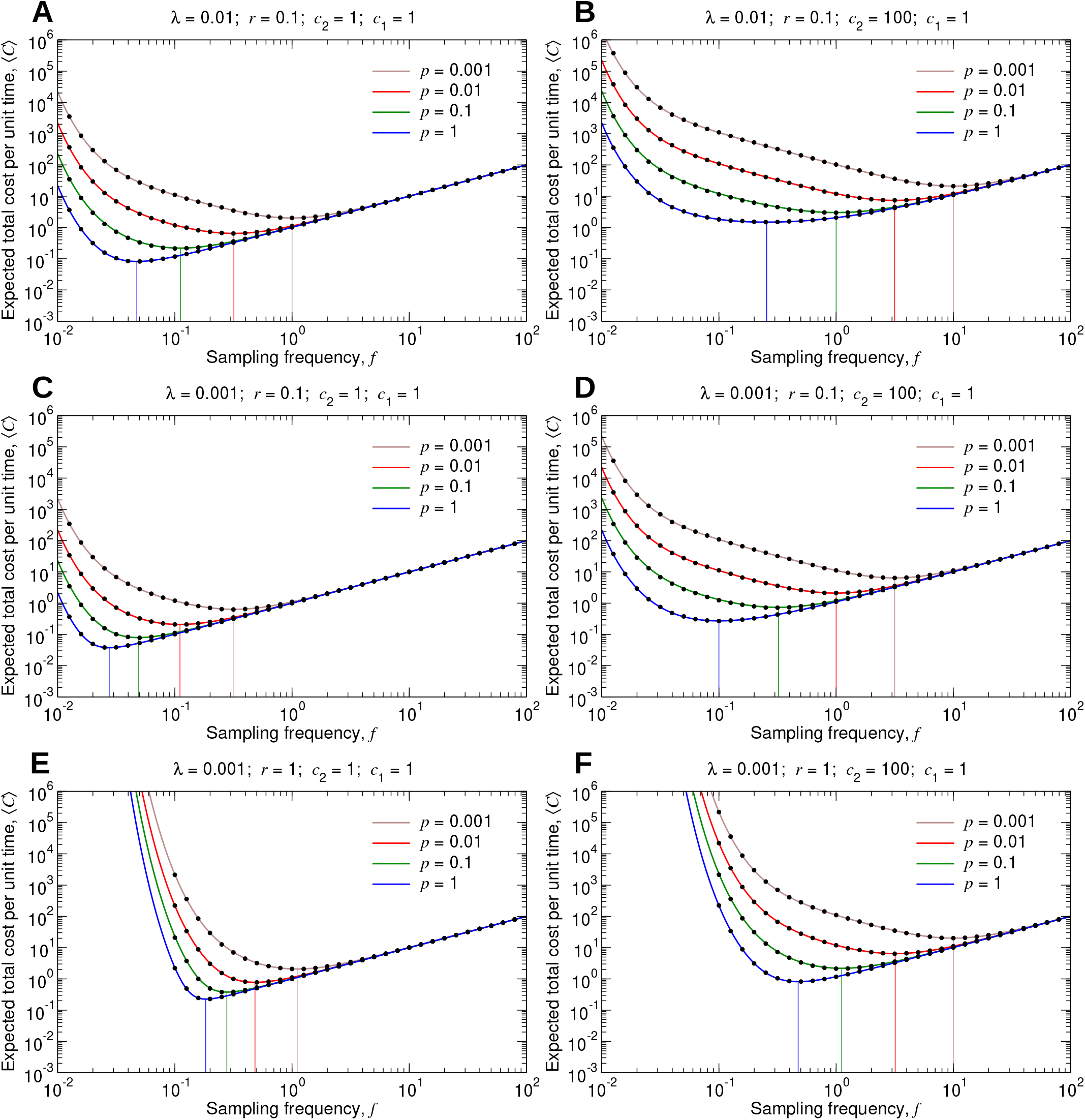
Expected total cost for a particular type of pathogen. (A through F) We set *c*_1_ = 1, and we plot *C*, given by Equation (1), as a function of *f* for several values of *λ, r, c*_2_, and *p*. The black dots are measurements of the expected total cost from simulating the true stochastic process. The 95% confidence intervals are smaller than the size of the data points. The vertical lines show the sampling frequencies for which the expected total cost is minimal in each case.

In designing and performing environmental surveillance, we do not know *a priori* the characteristics of a particular pathogen that may be introduced and result in an outbreak. Rather, for optimizing surveillance, the requirement is to have an understanding of the likely characteristics of new pathogens that might emerge. As a simple example, suppose that our surveillance platform is capable of detecting not just one but two different pathogens. Further, suppose that these two pathogens have different costs and are initiated at different rates. Let *c*_2_(1) denote the per-case cost for the first pathogen, and let *c*_2_(2) denote the per-case cost for the second pathogen. Similarly, let *λ*(1) denote the rate of introductions for the first pathogen, and let *λ*(2) denote the rate of introductions for the second pathogen. For this scenario, the expected infection cost per unit time is equal to [*λ*(1)][*c*_2_(1)][⟨N ⟩] + [*λ*(2)][*c*_2_(2)][⟨N ⟩]. It is also possible that the two pathogens differ in their growth rates and in their susceptibility to being detected. The first pathogen might have corresponding parameters *r*(1) and *p*(1), while the second pathogen might have parameters *r*(2) and *p*(2). As a result, the expected size of an outbreak of the first pathogen, ⟨N ⟩(1), might be different from the expected size of an outbreak of the second pathogen, ⟨N ⟩(2). The expected infection cost per unit time is then equal to [*λ*(1)][*c*_2_(1)][⟨N ⟩(1)] + [*λ*(2)][*c*_2_(2)] [⟨N ⟩(2)]. If there are more than two types of pathogens that can emerge and be detected by our surveillance platform, then in the calculation of the expected infection cost per unit time, we would simply add another term for each additional pathogen.

An important point is that the possible values of the parameters *c*_2_, *r*, and *p* that a pathogen can have are not discrete. Accordingly, let d*c*_2_ d*r* d*p λ*′(*c*_2_, *r, p*) denote the (infinitesimal) rate at which pathogens with per-case cost *c*_2_, growth rate *r*, and detection probability *p* emerge. In this more general treatment, *λ*′(*c*_2_, *r, p*) is a rate density that is a function of *c*_2_, *r*, and *p*. To calculate the expected pathogen cost, we integrate d*c*_2_ d*r* d*p λ*′(*c*_2_, *r, p*) *c*_2_⟨N ⟩ over all possible values of *c*_2_, *r*, and *p*. Accounting for all possible types of pathogens that might emerge, the expected total cost per unit time, ⟨C′⟩, is equal to

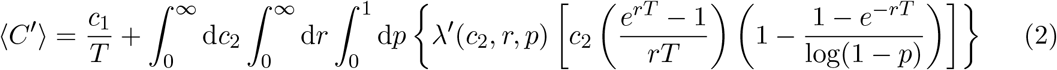

Equation (2) can be quickly calculated numerically for different values of the testing period, *T* . The value of 1*/T* for which the expected total cost is lowest specifies the optimal testing frequency, *F* ^*^. From Equation (2), we have

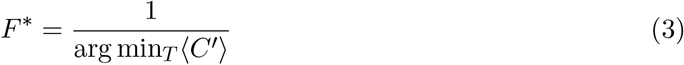

Figure 4 shows how this works. In Figure 4A, we show one possible form for the probability density function for *c*_2_. Pathogens with little or no associated cost (i.e., those for which *c*_2_ is close to zero) are most common, while more harmful pathogens occasionally arise. The parameter *a* controls the shape of the probability density function. For smaller values of *a*, the distribution has a longer tail, meaning that there is a higher chance that a new pathogen is harmful. In Figure 4B, we use this form for the probability density function for *c*_2_, we set *r* = 0.1 and *p* = 0.01, we set the total rate of emergence of new pathogens to 0.001, and we plot the expected total cost, ⟨*C*′ ⟩. (In the specification of *λ*′, *δ* denotes the Dirac delta function.) For smaller values of *a*, the optimal testing frequency for environmental surveillance increases accordingly.

**Figure 4:**
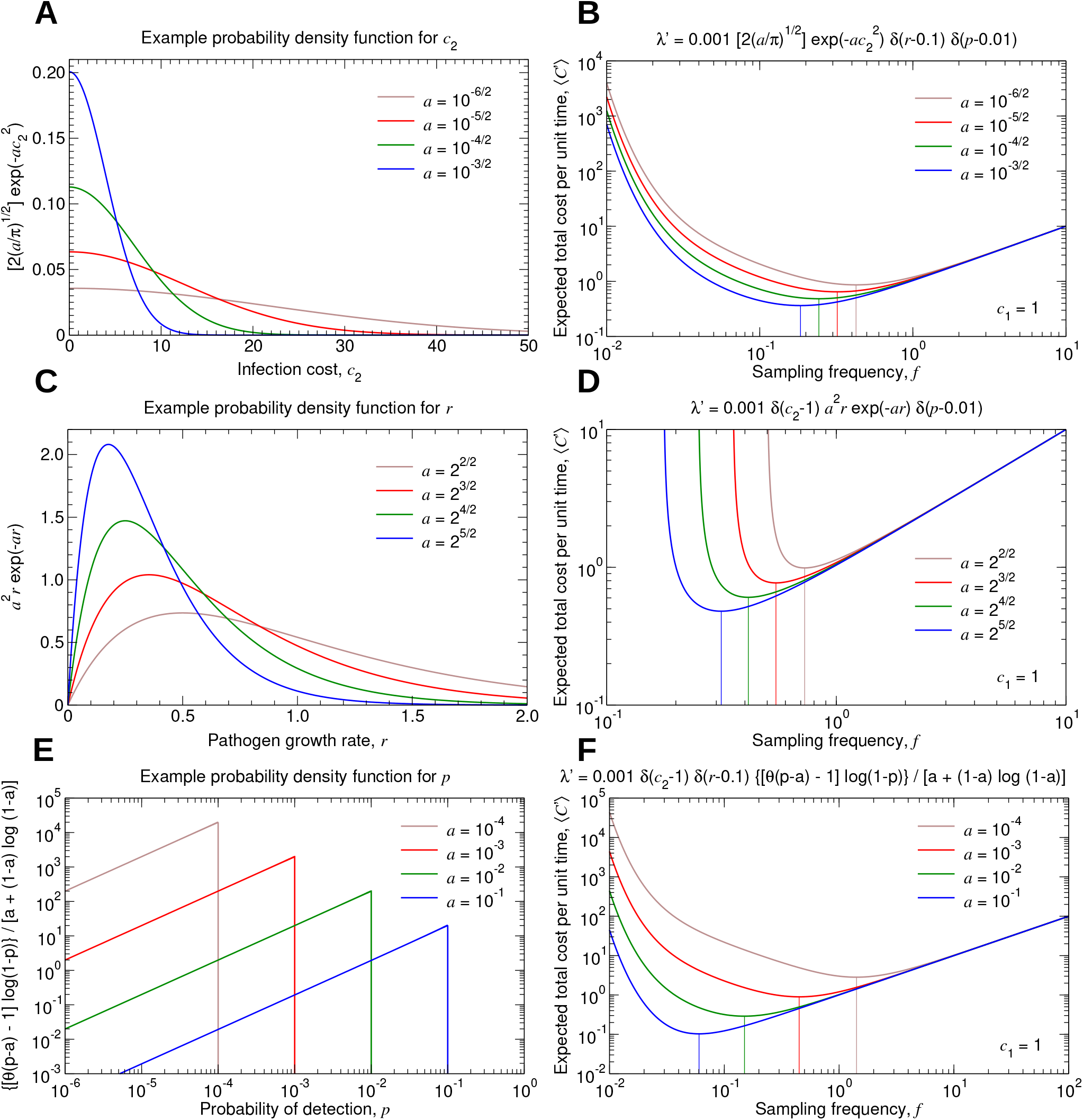
Expected total cost accounting for many types of pathogens. (A, C, and E) We show example probability density functions for the parameters *c*_2_, *r*, and *p*, respectively. For each case, we introduce a single parameter, *a*, which controls the shape of the probability density function. (B, D, and F) We set ⟨*c*_1_⟩ = 1, and we plot *C*′, given by Equation (2), as a function of *f* for several rate density functions, *λ*′(*c*_2_, *r, p*). The vertical lines show the sampling frequencies for which the expected total cost is minimal in each case.

Similarly, in Figure 4C, we show one possible form for the probability density function for *r*. We again use the parameter *a* to control the shape of the probability density function. Smaller values of *a* result in a longer tail to the distribution, so that pathogens with more rapid growth rates are more likely to arise. In Figure 4D, we use this form for the probability density function for *r*, we set *c*_2_ = 1 and *p* = 0.01, we set the total rate of emergence of new pathogens to 0.001, and we plot ⟨C′⟩. For smaller values of *a*, we must sample the environment more frequently. Notice that as the sampling frequency decreases below its optimum, the expected total cost rapidly increases. This is because there is a chance that pathogens with unusually large growth rates are introduced, and if their subsequent exponential growth is not halted soon enough, then the resulting pathogen-associated costs can become extremely large.

In Figure 4E, we show one possibility for the probability density function for *p*. For smaller values of *a*, there is a higher chance of pathogens being introduced that have low sensitivity to being detected. Using this form for the probability density function for *p* in Figure 4F, setting *c*_2_ = 1 and *r* = 0.1, and setting the total rate of emergence of new pathogens to 0.001, we plot ⟨C′⟩. (In the specification of *λ*′, *θ* denotes the Heaviside step function.) Smaller values of *a* result in a larger optimal testing frequency. Details on Figure 4 are provided in the Supplementary Information.

The example probability density functions in Figure 4 were chosen here for convenience: they have nice analytical forms, and they admit simple analytical solutions when substituted into Equation (2). For optimizing an environmental or vector surveillance system in practice, one would construct a estimated form for *λ*′(*c*_2_, *r, p*) based on experimental or observational data, and the optimal testing frequency would be determined numerically using Equation (3). Optimization of environmental or vector surveillance thus requires an understanding of the cost of each sampling and testing event, *c*_1_, and an understanding of the function for the rate of emergence of new pathogens, *λ*′(*c*_2_, *r, p*).

## Discussion

Equations (2) and (3) specify the optimal frequency at which to perform sampling and testing. Their use for optimizing testing frequency requires an estimation of the likely values of the per-case cost, *c*_2_, rate of growth, *r*, and susceptibility to detection, *p*, of any emerging pathogens. The rate density, *λ*′(*c*_2_, *r, p*), is large if pathogens with those parameter values are likely to emerge, and small otherwise. Estimating the dependence of *λ*′ on *p* entails many considerations. Molecular properties of emerging pathogens must be anticipated, and this must be interpreted in the context of whichever laboratory tests are used to detect it. Spatial structure of the landscape over which pathogens can emerge further influences the dependence of *λ*′ on *p*. For instance, if a pathogen emerges far from a wastewater treatment facility, then the number of infections in the vicinity of the location of sampling might be much smaller than the total size of the outbreak. This effect could be incorporated by using a reduced value of *λ*′. A similar consideration arises in sampling a vector population, where a pathogen might originate and begin spreading in individuals that are far from the nearest trap. Inferring the dependence of *λ*′ on *r* may be accomplished by analysis of historical data of either clinical cases or abundance of a pathogen in a vector species, together with maximum likelihood estimation. A larger number of outbreaks having occurred during a particular time period would correspond to a larger value of *λ*′. Optimization of testing frequency further requires a formal understanding of surveillance-related and pathogen-related costs [37, 38, 39]. Mathematically, the question of how to optimize a surveillance platform is undefined unless all relevant surveillance-related and pathogen-related costs are quantified in the same units. This is challenging, since the underlying factors are inherently very different in nature. Nonetheless, such understanding is essential if environmental and vector surveillance for infectious diseases is to be meaningfully optimized.

Our model for determining the optimal testing frequency is broadly applicable. If *α* is the probability of a test resulting in a false positive, then the sampling cost can be adjusted using the substitution *c*_1_ → *c*_1_ + *αk*, where *k* is the cost due to a false positive. This consideration can be extended by assuming that the probability of a false positive is dependent on *p*, i.e., *α* → *α*(*p*). Although the long-time dynamics of an emerging pathogen can show complex behavior, the early-time dynamics are often approximately exponential, and the associated disease-related costs at early times are expected to scale roughly linearly with the size of the outbreak. Both of these features are incorporated in our model. Once a pathogen is detected and an intervention is implemented, spread of the pathogen and its associated costs are not immediately halted, and this is accounted for by making the substitution *λ*′(*c*_2_, *r, p*) → *λ*′(*κc*_2_, *r, p*), where *κ <* 1. A further consideration is that intervention is unlikely to completely eliminate the pathogen. Subsequent sampling and testing would then monitor for when the pathogen becomes sufficiently abundant again that additional intervention is warranted. This may be approximately described by using a rate density for introductions that is time-dependent (i.e., *λ*′(*c*_2_, *r, p*) → *λ*′(*c*_2_, *r, p, t*)) and increases if there was a recently suppressed outbreak. The increased value of *λ*′ accounts for the possibility of a follow-up outbreak due to cases that the intervention failed to extinguish. Changes in weather and climate affect the risk of an outbreak—especially for many vector-borne pathogens [40]—and this could also be modeled through time-dependence of *λ*′.

Our approach can be applied to answer another important question: Where should environmental sampling be performed? This would work by introducing a spatial structure in the model. The pathogen can be introduced in one location and then migrate to different locations as it proliferates. By numerically running the stochastic dynamics with spatial structure, an expected total surveillance and disease cost can be calculated. By trying different sampling locations, it is possible to find the sampling locations for which the expected total cost is minimal.

Environmental and vector surveillance are equally instrumental for tracking the prevalence of a pathogen [41]. An understanding of how and when to intervene is therefore essential [42, 43]. If false positives are too frequent, then intervention costs will accumulate, leading to costly surveillance. If the designated signal that is required for intervention is too strong, then the pathogen can spread to the point where intervention has limited effectiveness in mitigating disease-related costs. The optimal testing frequency could also be adjusted as new data become available [44]. If tracking indicates increased prevalence of a pathogen, then more frequent sampling and testing might be warranted. A further possibility is that testing could be performed frequently over several seasons to gain an understanding of the typical seasonal behavior for new pathogens or in new ecological settings to inform optimization and more efficiently track the abundance of the pathogen.

Our model and its many possible extensions can thus inform the design of these critical aspects of environmental and vector surveillance platforms. Our work provides a general and robust foundation for mechanistic optimization of environmental surveillance for infectious diseases.

## Data Availability

All code for running the simulations and reproducing the figures is available at https://github.com/jolejarz.

## Acknowledgments

This project has been funded in part by contract 200-2016-91779 with the Centers for Disease Control and Prevention. Disclaimer: The findings, conclusions, and views expressed are those of the author(s) and do not necessarily represent the official position of the Centers for Disease Control and Prevention (CDC). S.M.K. received funding from NIH T32 training grant 2 T32 AI 7535-21 A1. M.L. is grateful for funding from the Morris-Singer Fund.

## Competing interests

The authors declare no competing interests.

## Supplementary Information

This Supplementary Information is organized as follows: In Section 1, we describe the surveillance protocol under consideration, and we derive the surveillance cost per unit time. In Section 2, we define the process by which new pathogens emerge, we define the dynamics of a pathogen that is growing in abundance, we define the manner in which each pathogen is detected, and we calculate the expected infection cost per unit time. In Section 3, we calculate the expected total cost per unit time, and we determine the optimal testing frequency. In Section 4, we describe how our model generalizes to account for the emergence of pathogens with different characteristics.

### 1 Surveillance cost

An important consideration for implementing environmental surveillance for pathogens is the frequency at which tests are performed. Environmental sampling and testing should be done frequently enough that an emerging pathogen is intercepted quickly, but not so frequently that surveillance costs outweigh the benefits of early detection. Here, we assume that whenever the environment is sampled and a test is conducted, a surveillance cost equal to *c*_1_ is incurred. We also assume that surveillance costs are additive, so that if *n* tests are performed, then the total surveillance cost is equal to *nc*_1_.

We consider that environmental tests are performed with period *T* . Since the cost of a single test is equal to *c*_1_, and since the time between tests is equal to *T*, the surveillance cost per unit time is given by

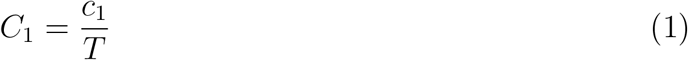

### 2 Expected infection cost

The costs incurred from the surveillance program itself must be considered in the context of infection-related costs. The costs due to an outbreak can vary depending on several factors:

- When the pathogen first appears in relation to the first environmental test that is performed following its introduction
- How the pathogen grows after it is introduced
- The sensitivity of the environmental testing program for detecting the pathogen
- The per-case infection cost

In this section, we describe each of these points in detail. Considering the full stochastic dynamics of pathogen initiation, pathogen growth, and pathogen detection, we derive a solution for the expected size of an outbreak when it is detected. We then derive an approximation for the expected size of an outbreak by assuming deterministic growth of a pathogen after it is initiated.

#### 2.1 Emergence of a pathogen

For determining the optimal testing frequency, we require knowledge of how new pathogens are introduced. We assume that the introduction of new pathogens follows a Poisson process. New pathogens are initiated independently and continuously in time at rate *λ*.

#### 2.2 Growth of a pathogen

We also require knowledge of how a pathogen increases in abundance once it first appears. Here, we assume that each instance of the pathogen makes new instances of the pathogen at rate *r* according to a Poisson process. Let *x*_*m,n*_(*t*) denote the probability that there are *n* copies of the pathogen at time *t*, given that there are *m* copies of the pathogen at time 0. In this section, we present the steps for calculating *x*_*m,n*_(*t*), beginning with the simplest cases and then progressing to the solution for any values of *m* and *n*, where *m ≤ n*.

##### 2.2.1 *m* = 1, *n* = 1

Suppose we start with a single instance of the pathogen at time 0 (*m* = 1). *x*_1,1_(*t*) gives the probability that the original instance of the pathogen has not produced any new instances of the pathogen up to time *t* (*n* = 1). *x*_1,1_(*t*) is given by

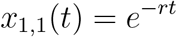

##### 2.2.2 *m* = 1, *n* = 2

Next, consider *x*_1,2_(*t*), which is the probability that the original instance of the pathogen has produced a single new instance of the pathogen by time *t* (*n* = 2). For this to occur, three things must happen: The original instance of the pathogen does not make any new instances of the pathogen between times 0 and *t*_1_, the original instance of the pathogen makes a new copy of itself at time *t*_1_, and neither of the two resulting instances of the pathogen make any new instances of the pathogen between times *t*_1_ and *t*. We must integrate over all values of *t*_1_ between 0 and *t*:

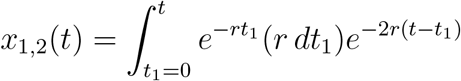

Simplifying, we have

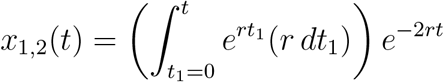

Performing the integration, we get

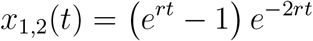

This then becomes

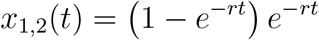

##### 2.2.3 *m* = 1, *n* = 3

Next, consider *x*_1,3_(*t*), which is the probability that the original instance of the pathogen has led to two new instances of the pathogen by time *t* (*n* = 3). For this to occur, five things must happen: The original instance of the pathogen does not make any new instances of the pathogen between times 0 and *t*_*2*_, the original instance of the pathogen makes a new copy of itself at time *t*_*2*_, neither of the two resulting instances of the pathogen make any new instances of the pathogen between times *t*_*2*_ and *t*_*1*_, one of the two instances of the pathogen makes a new copy of itself at time *t*_*1*_, and none of the three resulting instances of the pathogen make any new instances of the pathogen between times *t*_*1*_ and *t*. We must integrate over all values of *t*_*2*_ between 0 and *t*_*1*_, and we must integrate over all values of *t*_*1*_ between 0 and *t*:

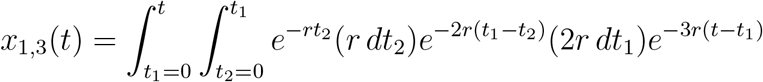

We can extend the range of the integration over *t*_*2*_ from *t*_*2*_ = 0 to *t*_*2*_ = *t* if we also divide by 2:

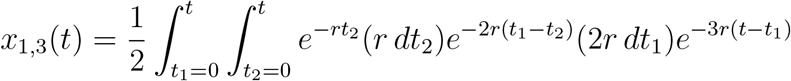

Simplifying, we have

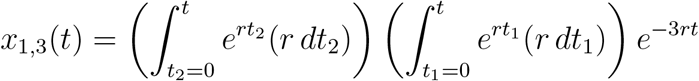

Performing the integration, we get

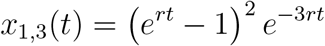

This then becomes

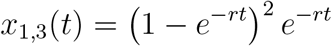

##### 2.2.4 *m* = 1, *n* = 4

Next, consider *x*_1,4_ (*t*), which is the probability that the original instance of the pathogen has led to three new instances of the pathogen by time *t* (*n* = 4). For this to occur, seven things must happen: The original instance of the pathogen does not make any new instances of the pathogen between times 0 and *t*_*3*_, the original instance of the pathogen makes a new copy of itself at time *t*_*3*_, neither of the two resulting instances of the pathogen make any new instances of the pathogen between times *t*_*3*_ and *t*_*2*_, one of the two instances of the pathogen makes a new copy of itself at time *t*_*2*_, none of the three resulting instances of the pathogen make any new instances of the pathogen between times *t*_*2*_ and *t*_*1*_, one of the three instances of the pathogen makes a new copy of itself at time *t*_*1*_, and none of the four resulting instances of the pathogen make any new instances of the pathogen between times *t*_*1*_ and *t*. We must integrate over all values of *t*_*3*_ between 0 and *t*_*2*_, we must integrate over all values of *t*_*2*_ between 0 and *t*_*1*_, and we must integrate over all values of *t*_*1*_ between 0 and *t*:

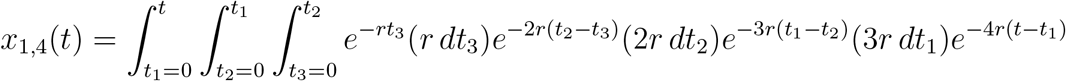

We can extend the range of the integration over *t*_*3*_ from *t*_*3*_ = 0 to *t*_*3*_ = *t* and the range of the integration over *t*_*2*_ from *t*_*2*_ = 0 to *t*_*2*_ = *t* if we also divide by 3!:

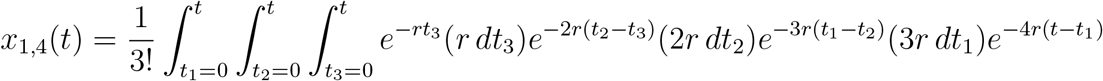

Simplifying, we have

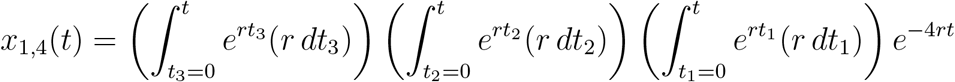

Performing the integration, we get

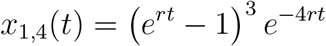

This then becomes

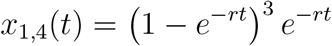

##### 2.2.5 *m* = 1, **any** *n*

We can generalize the calculation to arbitrary values of *n*:

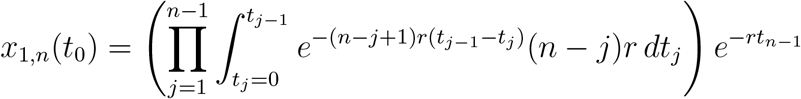

Changing the integration limits, we have

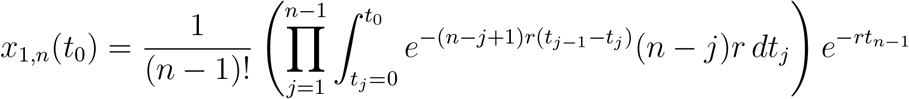

This simplifies to

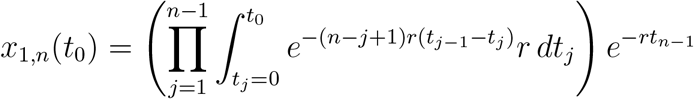

This becomes

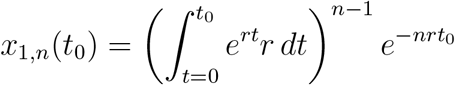

Performing the integration, we get

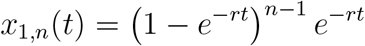

##### 2.2.6 Any *m* and *n*

Following the same procedure, we can calculate *x*_*m,n*_(*t*):

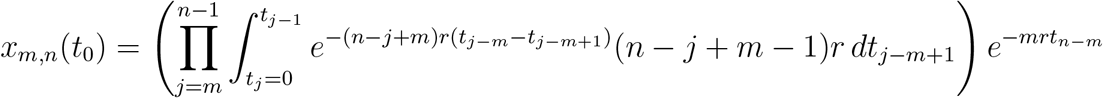

Changing the integration limits, we have

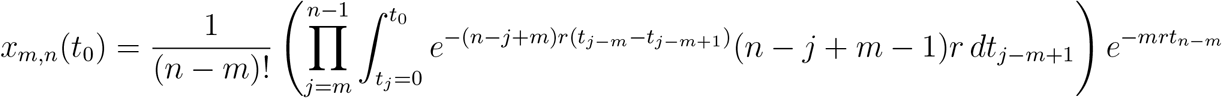

This simplifies further:

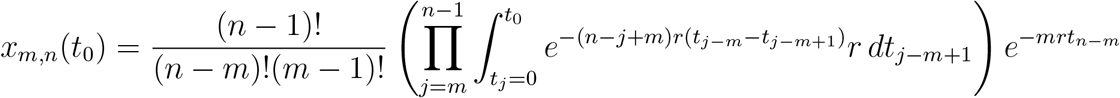

We can rewrite this as

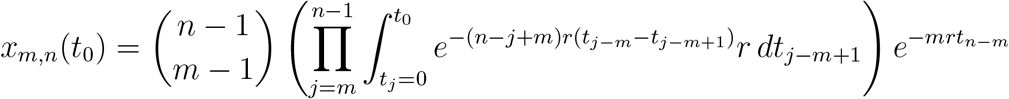

This becomes

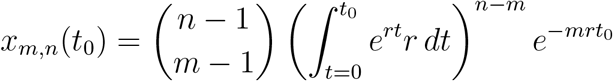

Performing the integration, we get

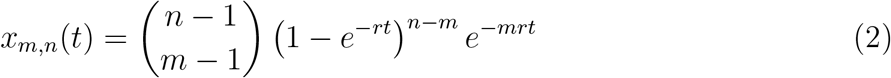

#### 2.3 Detection of a pathogen

We further require an understanding of how the outbreak is detected. Consider that there are *n* instances of the pathogen within a particular lineage when the environment is tested. We assume that each instance of the pathogen is not detected independently with probability *q*. The outbreak is not detected if and only if no instance of the pathogen is detected, which occurs with probability *q*^*n*^. Therefore, the pathogen is detected with probability 1 − *q*^*n*^.

We further assume that each lineage of the pathogen is detected independently of any other lineage. For example, suppose that two lineages of the pathogen are simultaneously present. Suppose that when the environment is tested, Lineage A contains *n*_*A*_ copies of the pathogen, and Lineage B contains *n*_*B*_ copies of the pathogen. In this case, Lineage A is detected with probability 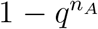, and Lineage B is detected with probability 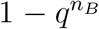. (If the rate of introduction of new pathogens, *λ*, is small, then simultaneous presence of two lineages would be a rare occurrence. Nonetheless, we describe this possibility so that the stochastic dynamics of pathogen initiation, pathogen growth, and pathogen detection are completely specified.)

#### 2.4 Expected size of an outbreak when it is detected

Using the stochastic rules presented above, and using Equation (2), we can derive a formula for the expected size of an outbreak when the pathogen is detected. For understanding the steps of the calculation, we define *X*_*i*_(*a*_*i*_) to be the probability that there are *i* testing events following the appearance of the pathogen that fail to detect the pathogen, and that there are *a*_*i*_ infections when the pathogen is detected.

We first consider the following question: What is the probability that the pathogen is detected in the first test following its appearance and that there are *a*_*0*_ instances of the pathogen when it is detected. This probability, which we denote *X*_*0*_(*a*_*0*_), is given by

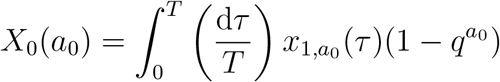

There are three components to this calculation:

- The pathogen is initiated at time *τ* before the testing event that detects it occurs. If the pathogen emerges just before the test that detects it is performed, then *τ* is slightly greater than 0. If the pathogen emerges just after the previous test, then *τ* is slightly less that *T* . Therefore, we have 0 *≤ τ < T* . Since new lineages appear independently and continuously in time, *τ* is equiprobably distributed between 0 and *T*, hence the integration 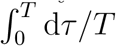.
- The pathogen begins as a single infection, and it grows to *a*_*0*_ infections at time *τ* since its appearance with probability 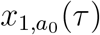.
- At least one of the *a*_*0*_ infections is detected with probability 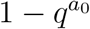.

Next, we can ask: What is the probability that the pathogen is detected in the second test following its appearance and that there are *a*_*1*_ instances of the pathogen when it is detected. This probability, which we denote *X*_*1*_(*a*_*1*_), is given by

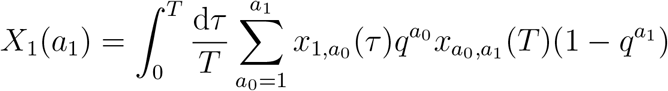

This calculation is understood as follows: The first test occurs at time *τ* after the pathogen appears, the pathogen grows to *a*_*0*_ infections at time *τ* after its emergence, none of those *a*_*0*_ infections are detected in the first test, the pathogen then grows to *a*_*1*_ infections at time *τ* + *T* after its emergence, and at least one of those *a*_*1*_ infections is detected in the second test. (We must sum over all values of *a*_*0*_ between 1 and *a*_*1*_.)

We can further ask: What is the probability that the pathogen is detected in the third test following its appearance and that there are *a*_*2*_ instances of the pathogen when it is detected. This probability, which we denote *X*_*2*_(*a*_*2*_), is given by

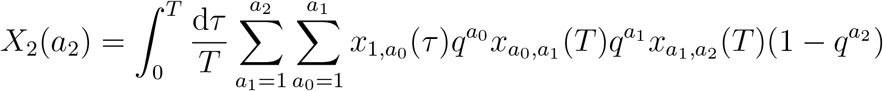

This calculation is understood as follows: The first test occurs at time *τ* after the pathogen appears, the pathogen grows to *a*_*0*_ infections at time *τ* after its emergence, none of those *a*_*0*_ infections are detected in the first test, the pathogen then grows to *a*_*1*_ infections at time *τ* + *T* after its emergence, none of those *a*_*1*_ infections are detected in the second test, the pathogen then grows to *a*_*2*_ infections at time *τ* + 2*T* after its emergence, and at least one of those *a*_*2*_ infections is detected in the third test. (We must sum over all values of *a*_*1*_ between 1 and *a*_*2*_ and over all values of *a*_*0*_ between 1 and *a*_*1*_.)

The calculation of *X*_*3*_(*a*_*3*_) follows in the same manner:

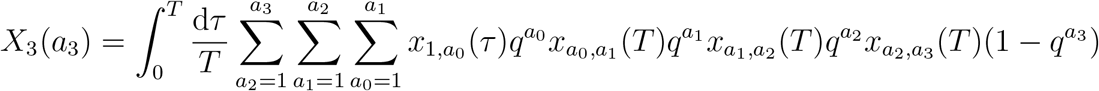

To calculate the expected size of an outbreak, we sum *X*_*m*_(*a*_*m*_)*a*_*m*_ over all possible numbers of failed tests (0 *≤ m < ∞*) and over all possible sizes of the outbreak when the pathogen is detected (1 *≤ a*_*m*_ *< ∞*):

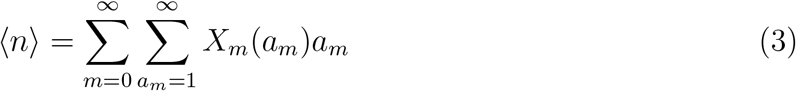

Equation (3) can be alternatively written as follows:

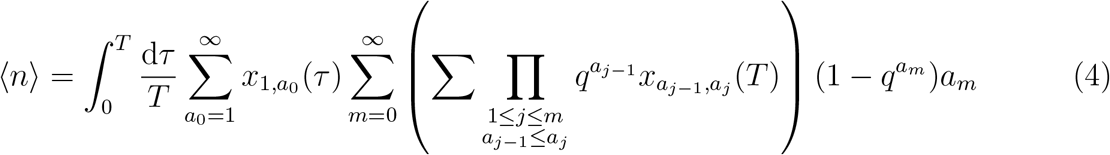

#### 2.5 Approximation for *⟨n⟩*

Equation (4) is analytically unwieldy. To make progress, we derive an approximate solution for the expected size of an outbreak by assuming that the pathogen grows deterministically after it is initiated (Figure S1). The first several steps of this process are as follows:

- The first infection occurs, and at time *τ* after its emergence, a test is performed. The size of the outbreak when the first test is performed is equal to *e*^*rτ*^
- If the pathogen is not detected in the first test, which occurs with probability 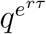, then the pathogen grows until the second test is performed, and the amount of growth of the pathogen between the first and second tests is equal to *e*^*r*(*τ*+*T*^ *− e*^*rτ*^ .
- If the pathogen is not detected in the first test and the second test, which occurs with probability 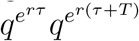, then the pathogen grows until the third test is performed, and the amount of growth of the pathogen between the second and third tests is equal to *e*^*r*(*τ* +2*T* )^ *− e*^*r*(*τ* +*T*)^.
- If the pathogen is not detected in the first test, the second test, and the third test, which occurs with probability 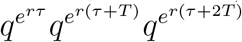, then the pathogen grows until the fourth test is performed, and the amount of growth of the pathogen between the third and fourth tests is equal to *e*^*r*(*τ*+3*T*^ *− e*^*r*(*τ*+2*T*^.

This process continues until the pathogen is detected. We therefore have the following result for the expected size of an outbreak:

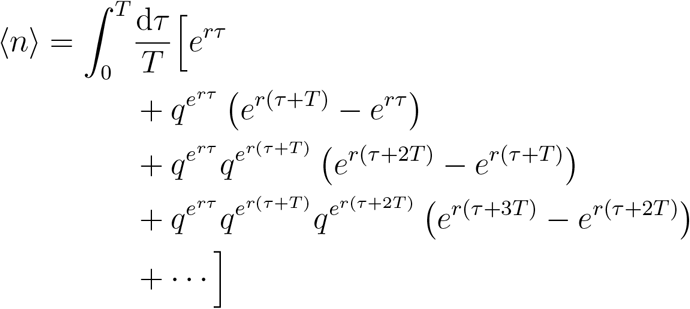

More compactly:

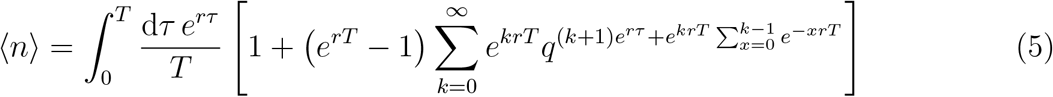

The process can also be considered by defining

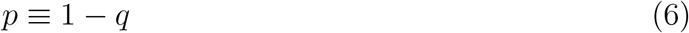

Here, *p* is the probability that a single infection is detected in a testing event, so that the probability that an outbreak of size *n* is detected in a testing event is given by 1 − (1 − *p*)^*n*^. Substituting Equation (6) into Equation (5), we have

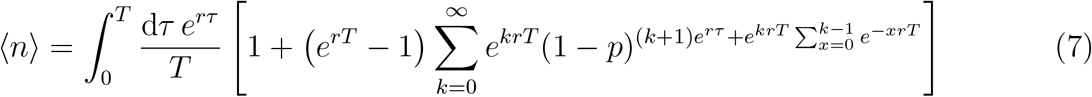

Next, we simplify Equation (7) in two limits: for the case *p* → 1 and for the case *p* → 0. We then construct an approximation for *⟨n⟩* for any value of *p*.

##### 2.5.2 *p* → 1

In the limit *p* → 1, Equation (7) simplifies:

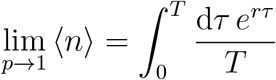

Performing the integration, we obtain

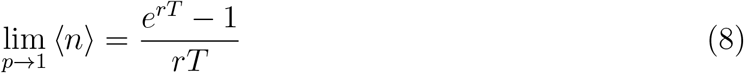

##### 2.5.2 *p* → 0

We can rewrite Equation (7) as follows:

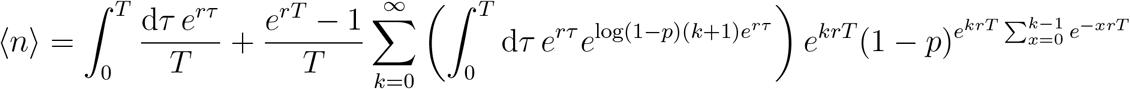

Performing the integration, this becomes

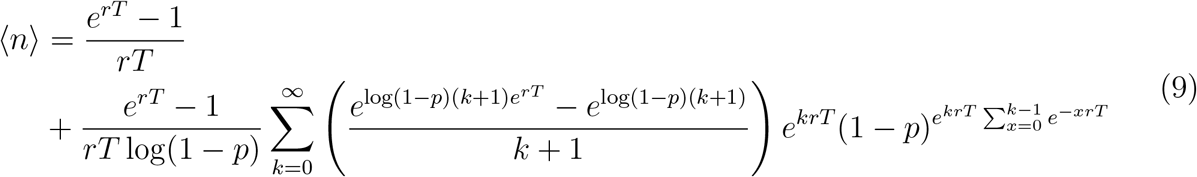

If *p* is small, then for an outbreak to be detected quickly, the testing period, *T*, must also be small. Considering that *p ≪* 1 and that *rT ≪* 1, the numerator of the expression in parentheses in Equation (9) can be approximated:

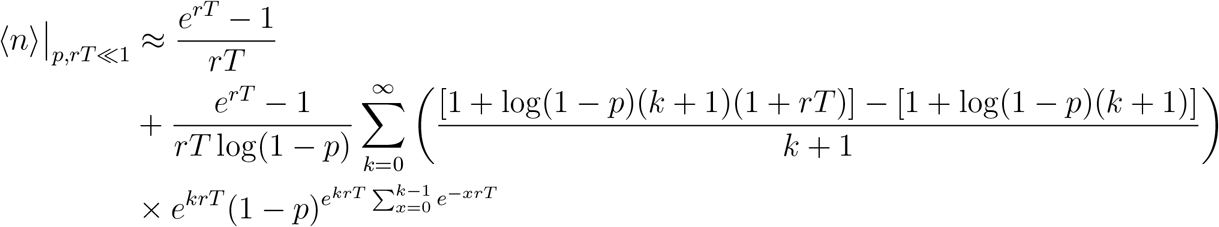

This becomes

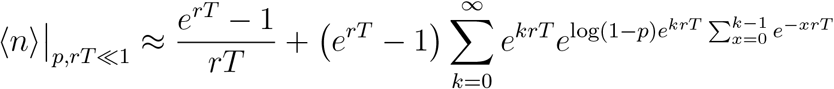

Next, we approximate 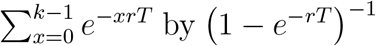, and we approximate the summation over *k* by an integration over *k*:

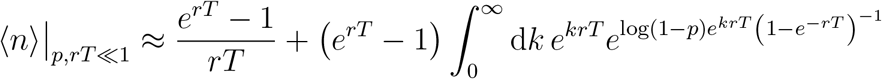

Performing the integration, this becomes

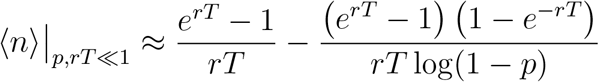

Simplifying, we have

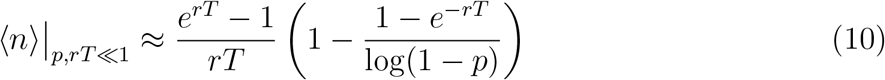

##### 2.5.3 0 *< p ≤* 1

Notice that in the limit *p* → 1, Equation (10) becomes equivalent to Equation (8). Therefore, for any value 0 *< p ≤* 1, we have the following approximate solution for *⟨n⟩*:

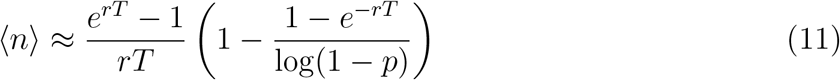

### 3 Expected total cost per unit time

For optimizing the testing frequency, the quantity of interest is the expected total cost per unit time. The surveillance cost per unit time, *C*_*1*_, is given by Equation (1). Let *⟨C*_*2*_*⟩* denote an approximation for the expected infection cost per unit time, and let *⟨C⟩* denote an approximation for the expected total cost per unit time. We have

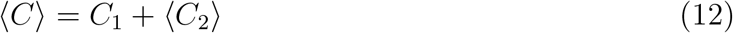

For determining ⟨*C*_*2*_⟩, we assume that each infection contributes a cost *c*_*2*_. If new lineages appear at rate *λ*, then *⟨C*_*2*_*⟩* is given by

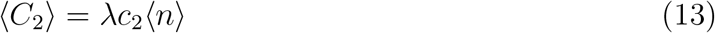

Substituting Equations (1), (13), and (11) into Equation (12), we obtain

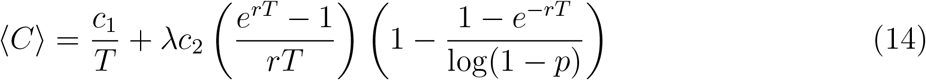

#### 3.1 Optimal testing frequency

Equation (14) specifies the expected total surveillance and pathogen cost. *⟨C⟩* is a function of the testing period, *T*, and we seek the value of *T* for which *⟨C⟩* is minimal. The first step is to show that *⟨C⟩* has a single minimum at a particular value of *T* . To do this, we differentiate *⟨C⟩* twice with respect to *T* :

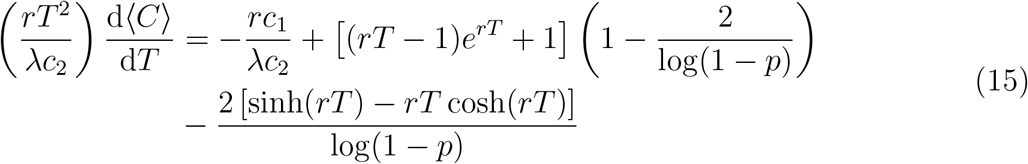

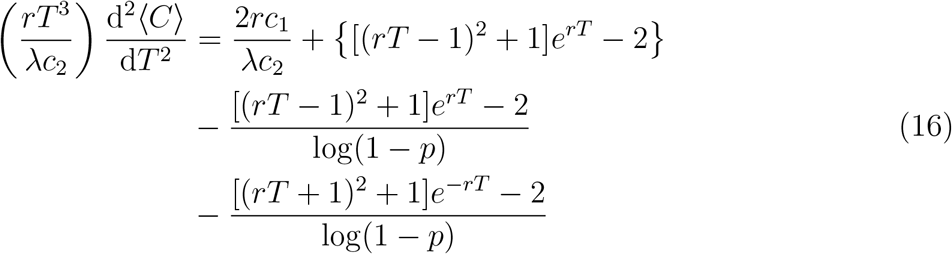

In Equation (16), the quantity *rT* ^*3*^*/*(*λc*_*2*_) is necessarily positive. If the right-hand side of Equation (16) is positive for positive values of *T*, then d^*2*^*⟨C⟩/*d*T* ^*2*^ is necessarily positive. Note that

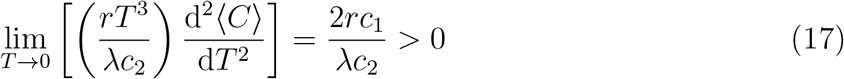

We also have

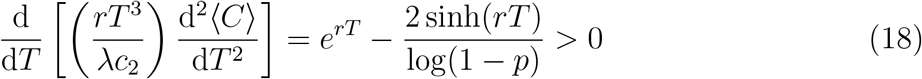

From Equations (17) and (18), it follows that the right-hand side of Equation (16) is necessarily positive. Therefore,

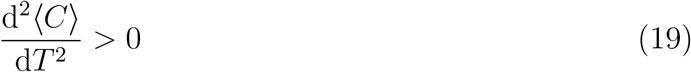

Next, note that

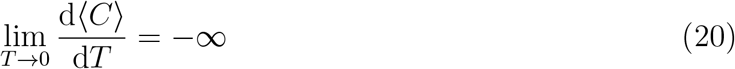

We also have

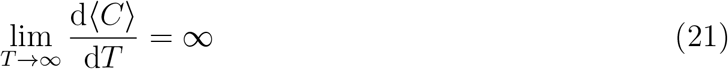

From Equations (20), (21), and (19), it follows that there is a single value of *T* for which *⟨C⟩* is minimized.

To determine the optimal testing period, we set d *⟨C⟩ /*d*T* = 0 and *T* = *T* ^*\**^ in Equation (15). We arrive at an implicit solution for the optimal testing period, *T* ^*\**^:

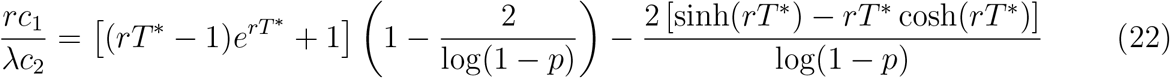

The optimal testing frequency is given by

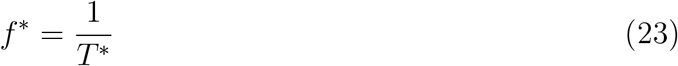

##### 3.1.1 Asymptotic behavior as *p* → 1

Taking the limit *p* → 1 in Equation (22), we obtain the following equation for *T*^*\**^:

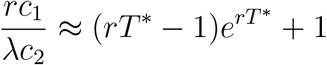

Letting *W*_*0*_(*x*) denote the principal branch of the Lambert W function, and using Equation (23), we obtain an explicit approximation for the optimal testing frequency:

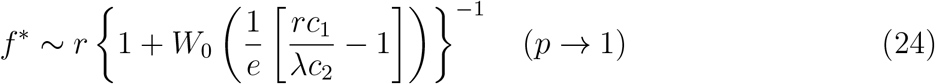

In Figure S2, we plot Equation (24) as a function of *c*_*2*_ for several sets of parameter values. We also plot measurements of the optimal testing frequency from simulating the true stochastic process.

##### 3.1.2 Asymptotic behavior as *p* → 0

For small values of *p*, the optimal testing frequency, *T* ^*\**^, is also small. To determine *T* ^*\**^, we consider that *p ≪* 1 and that *rT* ^*\**^ *≪* 1 in Equation (22). We use the approximations 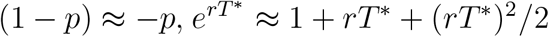, sinh(*rT* ) *≈ rT* + (*rT*^*\**^)^3^*/*3!, and cosh(*rT* ^*\**^) *≈* 1 + (*rT* ^*\**^)^*2*^*/*2:

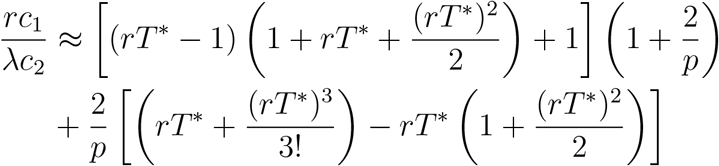

Simplifying, we have

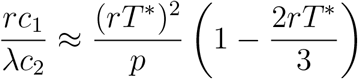

For small values of *rT* ^*\**^, the second term on the right-hand side is negligible relative to the first. Using Equation (23), we solve approximately for the optimal testing frequency:

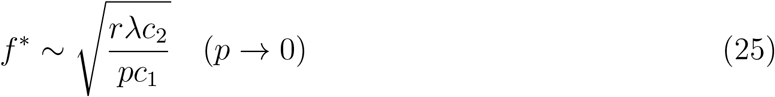

In Figure S3, we plot *f* ^*\**^ from Equations (22) and (23) as a function of *p* for several sets of parameter values. We also plot measurements of the optimal testing frequency from simulating the true stochastic process. For small values of *p, f* ^*\**^ is approximately given by Equation (25).

### 4 Distribution of pathogen-related parameters

The calculation of the expected infection cost per unit time, *(C*_*2*_), assumes that, for each lineage that appears, the pathogen-specific parameters *c*_*2*_, *r*, and *p* are the same. The expected infection cost per unit time is then just the expected cost due to a single lineage multiplied by the rate, *λ*, at which those lineages arise.

More generally, we can consider d*c*_*2*_ d*r* d*p λ*′(*c*_*2*_, *r, p*) to be the (infinitesimal) rate at which lineages with pathogen-specific parameters *c*_*2*_, *r*, and *p* appear. In this generalized model, let 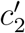 denote an approximation for the expected infection cost per unit time, and let ⟨*C*′⟩ denote an approximation for the expected total cost per unit time. We have

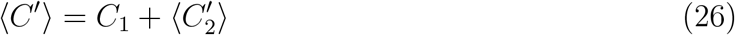

With knowledge of the rate density function, *λ*′(*c*_*2*_, *r, p*), we are able to compute the expected infection cost per unit time by integrating over all possible values of *c*_*2*_, *r*, and *p*:

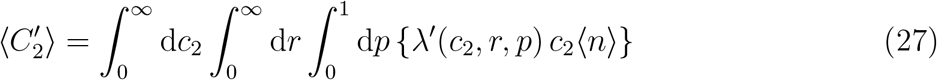

Substituting Equations (1) and (27) into Equation (26), we obtain

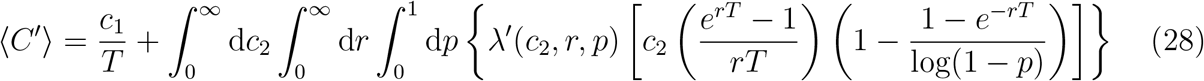

The optimal testing frequency is given by

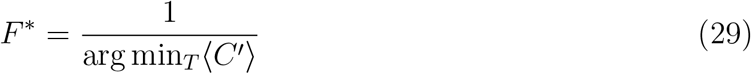

Equations (28) and (29) can be solved numerically to determine the optimal testing frequency. Below, we consider several simple examples for which Equation (28) can be solved analytically to show how the model works.

#### 4.1 Example 1

As the simplest example of using Equation (28), consider that only a single type of pathogen can emerge. The pathogen has per-case cost 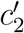, growth rate *r*′, and probability of detection *p*′, and new lineages are introduced at rate *λ*. The rate density function, *λ*′(*c*_*2*_, *r, p*), is given by

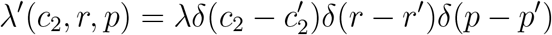

Here, *δ* denotes the Dirac delta function. When this form for *λ*′(*c*_*2*_, *r, p*) is substituted into Equation (28) and the integrations over *c*_*2*_, *r*, and *p* are performed, we obtain

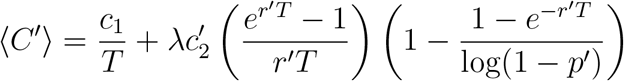

Thus, Equation (28) reduces to Equation (14) for the case where only a single type of pathogen with fixed parameters can emerge.

#### 4.2 Example 2

Next, consider the possibility that two different types of pathogens can emerge. Pathogen 1 has parameters 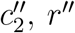, *r*′, and *p*′, while Pathogen 2 has parameters 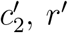, and *p*^*″*^. Lineages of Pathogen 1 are introduced at rate *λ*_*1*_, and lineages of Pathogen 2 are introduced at rate *λ*_*2*_. The corresponding rate density function is

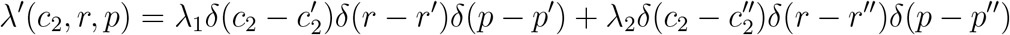

When this form for *λ*′(*c*_*2*_, *r, p*) is substituted into Equation (28) and the integrations are performed, we obtain

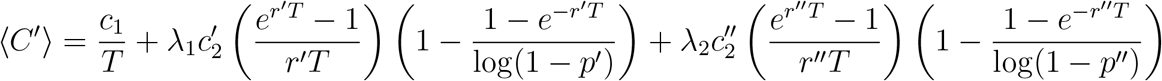

The expected total cost per unit time, ⟨*C*^*″*^ ⟩, is therefore equal to the surveillance cost per unit time, plus the expected infection cost per unit time for Pathogen 1, plus the expected infection cost per unit time for Pathogen 2.

#### 4.3 Example 3

These considerations can be extended to the case where many different types of pathogens can emerge. Let Pathogen *n* have per-case cost *c*_2,*n*_, growth rate *r*_*n*_, and probability of detection *p*_*n*_. The rate density function, *λ*^*″*^(*c*_*2*_, *r, p*) is given by

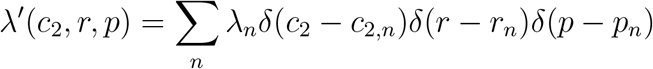

Substituting this into Equation (28) and integrating yields

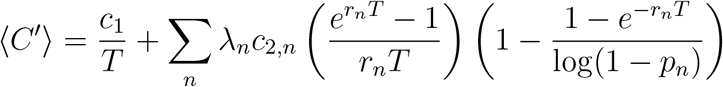

The expected infection cost is therefore linear—i.e., we add together the expected infection costs for each of the *n* possible types of pathogens, and this sum equals the total expected infection cost.

#### 4.4 Example 4

The possible parameter values that any new pathogen can have are not discrete. They are continuous. To show how this works, consider the following form for the rate density function:

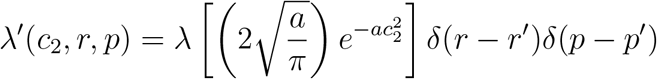

For this case, new pathogens have growth rate *r*^*″*^ and probability of detection *p*^*″*^. New pathogens can, however, have any real value of *c*_*2*_ that is nonnegative. For any lineage that is introduced, it’s value of *c*_*2*_ is most likely to be close to zero, while larger values of *c*_*2*_ occur more rarely. The parameter *a* controls with with of the probability density function for *c*_*2*_. For smaller values of *a*, this distribution has a longer tail, and the expected value of *c*_*2*_ for any new pathogen increases. Substituting this form for the rate density function into Equation (28) and integrating, we have

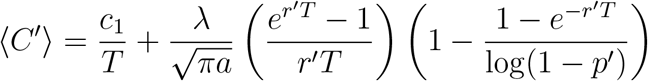

#### 4.5 Example 5

For this example, we suppose that any new pathogen has per-case infection cost 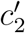 and probability of detection *p*^*″*^, while the growth rate, *r*, can be any nonnegative real number. We use the following form for the rate density function:

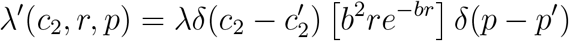

Substituting this into Equation (28) and integrating, we have

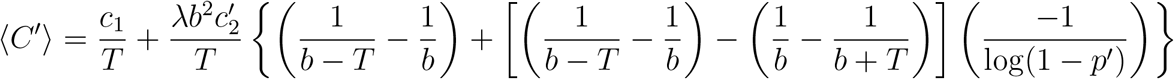

#### 4.6 Example 6

We can also model the case where new pathogens have per-case cost 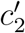 and growth rate *r*^*″*^, while the probability of detection, *p*, can be any real number between 0 and 1. Suppose that the rate density function has the following form:

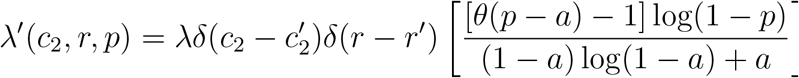

Here, *θ* denotes the Heaviside step function. Substituting this into Equation (28) and integrating, we obtain

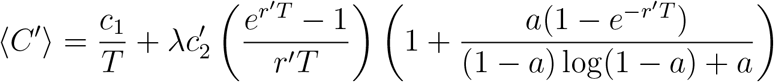

**Figure S1:**
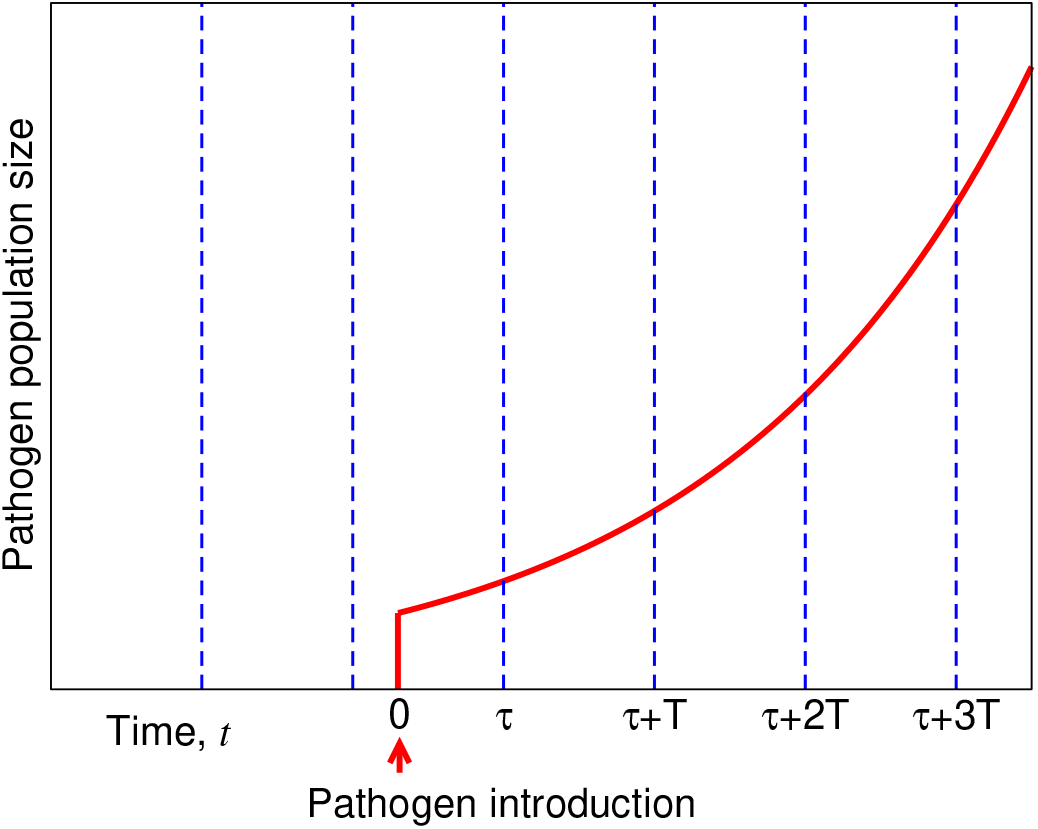
Schematic showing deterministic growth of the pathogen. For calculating an approximation for the expected size of an outbreak when it is detected, we can assume that the size of the outbreak grows deterministically.

**Figure S2:**
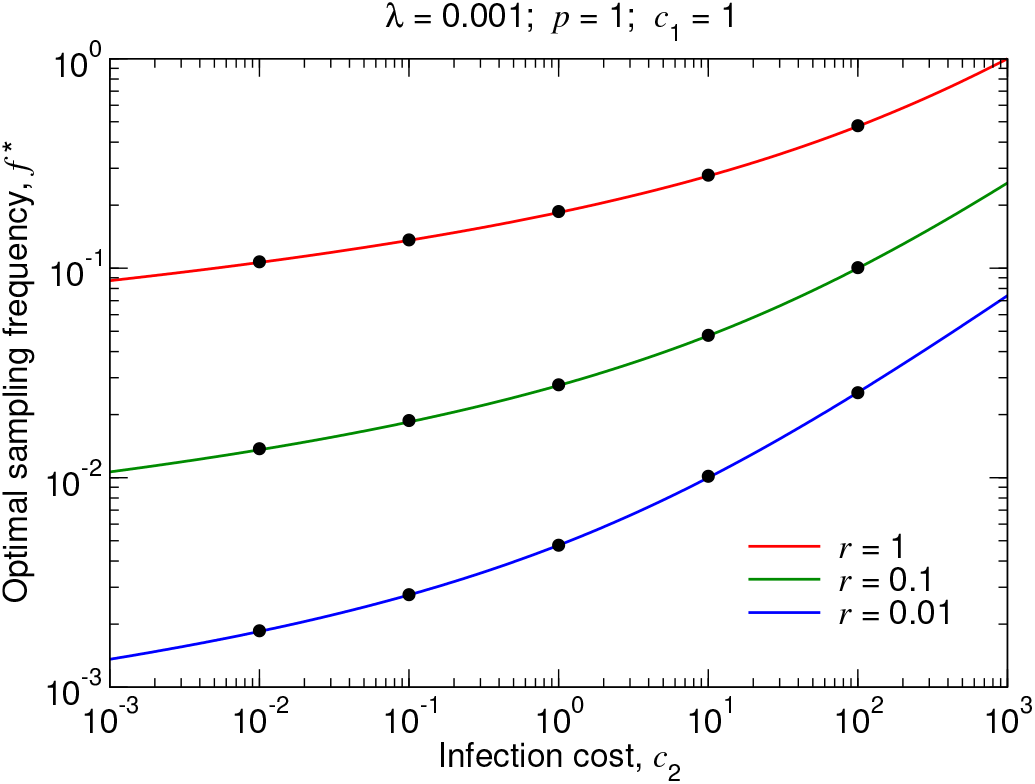
Optimal testing frequency for. *p* = 1. For different values of *r*, we plot the optimal testing frequency, *f* ^∗^, given by Equation (24), as a function of *c*_2_. The black dots are measurements of the optimal testing frequency from simulating the true stochastic process. The 95% confidence intervals are smaller than the size of the data points.

**Figure S3:**
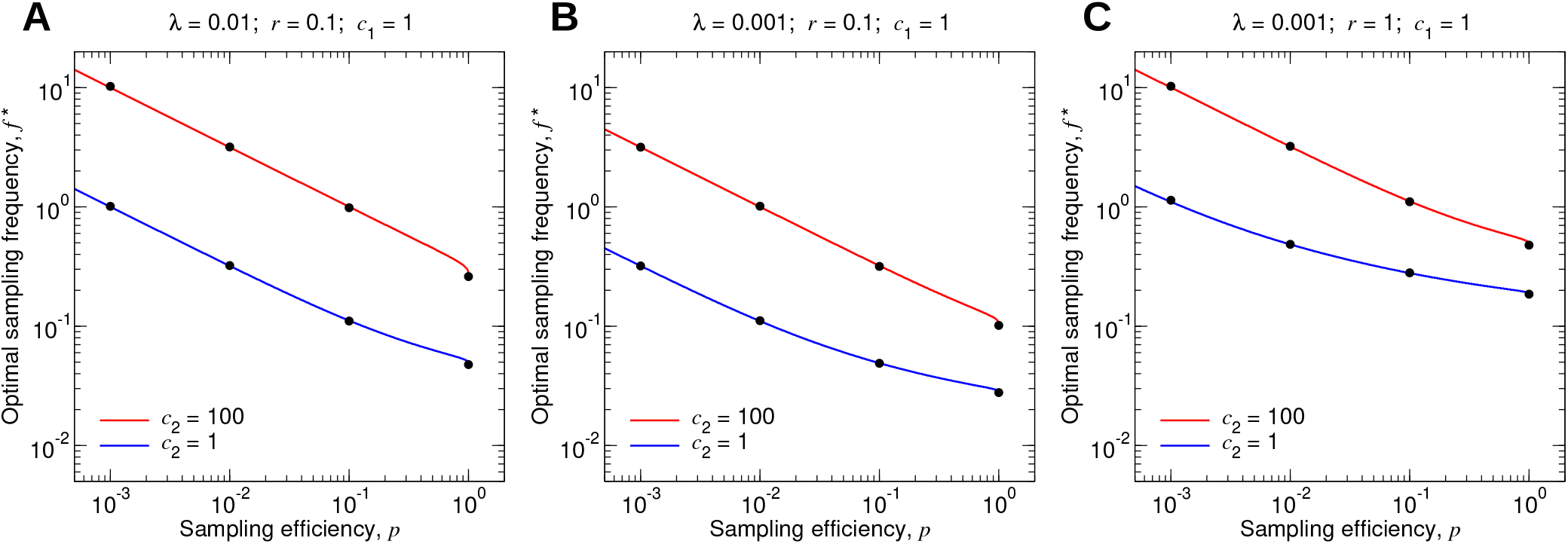
Optimal testing frequency for. *p* 1. For different values of *c*_2_, we plot the optimal testing frequency, *f* ^∗^, given by Equations (22) and (23), as a function of *p*. The black dots are measurements of the optimal testing frequency from simulating the true stochastic process. The 95% confidence intervals are smaller than the size of the data points.

## Notes

### Competing Interest Statement

The authors have declared no competing interest.

